# The Antipsychotic Load (AP_Load2026): a normalised prescribed-dose index for schizophrenia research

**DOI:** 10.64898/2026.11.09.26367598

**Authors:** Emilio Fernandez-Egea, Rajeev Krishnadas, Christopher Jenkins, Rashmi Patel, Rudolf N. Cardinal, Noham Wolpe

## Abstract

**Background and Hypothesis:** Schizophrenia research routinely reduces antipsychotic treatment to a single number, but the available summaries disagree. Chlorpromazine equivalents, the WHO Defined Daily Dose (DDD) and the percentage of the licensed maximum (%BNFmax) rank the same patient differently, most of all for clozapine, whose dose–concentration relationship varies several-fold with sex and smoking. We hypothesised that referencing clozapine dose to a target individualised for sex and smoking would track plasma clozapine better than dose-proportional measures.

**Study Design:** In the Antipsychotic Load (AP_Load2026), each daily dose is expressed as a percentage of a reference dose, summed without a cap. Clozapine is referenced to a target individualised for sex and cigarettes per day, under two published anchor schemes, and other antipsychotics to a dose reconciling the WHO DDD and the licensed maximum. The clozapine term was tested against plasma clozapine (1316 assessments, 255 patients) with log–log mixed-effects models, and the non-clozapine reference against independently derived published doses.

**Study Results:** The individualised reference tracked plasma clozapine better than any dose-proportional measure (marginal R² 0.249 v. 0.166; ΔR² 0.082, 95% CI 0.044–0.120). The scheme with the larger smoking correction fitted better (0.249 v. 0.231), an ordering robust to quality-control variants and co-prescription. Of five candidate references for other antipsychotics, %BNFmax was the least consistent with external criteria.

**Conclusions:** AP_Load2026 is a transparent framework for normalising prescribed antipsychotic dose. Its clozapine reference is validated against a biomarker, whereas the reference for other antipsychotics is a calibrated convention at least comparable with existing alternatives. The index is particularly suited to treatment-resistant and clozapine-enriched cohorts.

## Introduction

Most clinical studies in schizophrenia, whether observational or experimental, must reduce antipsychotic treatment to a single figure, whether to describe the sample or to adjust for medication as a potential confounder. Three broad approaches remain in wide use, and they can disagree substantially. Chlorpromazine or olanzapine equivalents, built on potency ratios,^1,2^ are used both for current dose^3,4^ and, accumulated over time as dose-years, for lifetime exposure.^5,6^ The WHO Defined Daily Dose (DDD), an administrative unit of drug utilisation,^7,8^ dominates register studies, usually as DDD/day bands.^9,10^ The percentage of the licensed British National Formulary (BNF) maximum (%BNFmax)^11,12^ offers near-universal coverage.

Each has a well-recognised weakness. Equivalence methods depend on potency ratios that are contested, source-dependent, and are not a single method: published schemes reference doses to chlorpromazine, olanzapine, risperidone or haloperidol and are derived variously from expert consensus, minimum effective doses or dose-response curves,^13–15^ with chlorpromazine and olanzapine equivalents the most widely reported. The DDD is an assumed average maintenance dose that can diverge from clinical potency.^8^ The %BNFmax denominator, though authoritative and comprehensive,^16^ is a safety ceiling rather than a marker of effective dose. In practice the choice of exposure proxy is seldom made explicit and is rarely justified.

Because these measures can rank the same patient very differently, findings expressed in one are difficult to compare or pool with findings expressed in another. Risperidone and clozapine are illustrative. A 6 mg risperidone dose is a substantial everyday dose (1.2 DDDs) yet only 37.5% of the 16 mg BNF maximum, so a measure anchored to that maximum understates it. Clozapine is more problematic still. Its licensed maximum (900 mg/day) and DDD (300 mg/day) are fixed numbers, but in clinical practice the effective dose is set by clearance, and therefore by sex and smoking, with typical targets ranging from about 250 to 550 mg/day.^17,18^ A male smoker on 450 mg/day sits at only half the licensed maximum yet at 1.5 DDDs. The same dose can therefore read as conservative, above average, or excessive depending solely on the chosen yardstick. Because clozapine is frequently combined with a second antipsychotic in treatment-resistant schizophrenia (TRS),^19,20^ these denominator errors compound under polypharmacy.

Here we aimed to define the Antipsychotic Dose Load (AP_Load2026), a normalised prescribed-dose index built on an explicit, documented and dated reference table. Its two components are constructed and evidenced differently. Clozapine is referenced to a target individualised for sex and smoking, and that reference can be tested against measured plasma clozapine. No such test exists for the other antipsychotics, since a co-prescribed drug does not alter clozapine concentrations; their reference is instead calibrated against published reference doses derived independently of both the DDD and the licensed maximum.

## Method

We divided the process into index definition and validation.

### Phase 1: definition of the index

For each antipsychotic the prescribed daily dose is expressed as a percentage of a reference dose; where more than one antipsychotic is prescribed, the percentages are added. The total is not capped, since values above 100% are meaningful and arise from high-dose treatment, polypharmacy or augmentation. Parallel loads for other psychotropic classes were also constructed by the same arithmetic, but they are not validated and are provided in the **Supplementary Material** as optional exploratory outputs only.

### Clozapine

Clozapine requires separate treatment. Because it is dosed to a target blood level and its clearance depends heavily on cytochrome P450 1A2 (CYP1A2) activity, a flat 900 mg/day denominator makes every clozapine patient appear lightly treated. We therefore reference clozapine to an individualised target, the dose expected to yield a plasma concentration of about 0.35 mg/L (350 ng/mL), modulated by sex and smoking status.^17,21^ Smoking enters continuously as cigarettes per day, with induction saturating at approximately ten per day. The inducing exposure is the polycyclic aromatic hydrocarbon content of tobacco smoke acting on hepatic CYP1A2 rather than nicotine itself, so e-cigarettes, nicotine replacement and heated-tobacco products do not induce the enzyme and their users are scored as non-smokers unless they also smoke tobacco. Notably, our anchors derive from predominantly European samples and a single UK validation cohort. Because clozapine clearance varies with ancestry, local recalibration may be needed wherever the necessary data exist.^17^

We used two candidate target schemes (**Table 1**). Rostami-Hodjegan et al^22^ reported a regression implying a smoking effect of about +62% on the required dose and a female effect of about −18%; the reference values given in the Maudsley Prescribing Guidelines^18^ are very similar. Reeves et al^23^ published a population-pharmacokinetic model giving 275 mg/day for a non-smoking man, +30% for smokers and −18% for women. We take from each only the sex- and smoking-stratified anchor doses, not the full model, and refer throughout to the Rostami-Hodjegan-derived and Reeves-derived schemes. The simplification bears more heavily on the Reeves-derived scheme, two of whose predictors, ethnicity and body weight, are not recorded in the validation extract.

**Table 1.** Clozapine individualised target dose (for ∼0.35 mg/L), mg/day, by sex and smoking under the two schemes. Smokers are shown at full induction; intermediate consumption is interpolated. Anchors are computed from the unrounded published values and rounded only for display. The Maudsley values are not shown separately, being near-identical to Rostami-Hodjegan.

|  | **Rostami-Hodjegan-derived** | **Reeves-derived** |
|---|---|---|
| Female, non-smoker | 265 | 226 |
| Male, non-smoker | 323 | 275 |
| Female, smoker | 431 | 293 |
| Male, smoker | 525 | 358 |

### Other antipsychotics

For non-clozapine agents we considered three candidate reference doses. The BNF maximum (%BNFmax) is transparent, current and available for almost every drug, but is a safety ceiling that often sits far above ordinary use. The DDD is closer to everyday dosing but is undefined for several newer agents. The third, which we adopt, is the geometric mean of the two, √(BNFmax × DDD), capped at 1.5 times the DDD. The geometric mean lies an equal proportional distance from each reference, so neither dominates by being the larger number, as it would under an arithmetic mean.

Two principles define the implementation. First, the cap applies to the reference dose in milligrams, not to the percentage it produces: risperidone’s reference becomes 7.5 mg, so a 6 mg dose scores 80%. In percentage terms the score for each drug is max(√(%BNFmax × %DDD), %DDD ÷ 1.5); capping the DDD-based percentage at 150% is a different operation and yields a lower figure. Second, the reconciliation is applied to each drug separately and the regimen score is the sum of the per-drug percentages, never a geometric mean of summed values. Worked examples are given in Table 5 and in the code repository (see Data availability). Agents without a DDD keep their BNF or national-label maximum. Long-acting injectables are referenced to their licensed formulation maximum per injection interval.^24^ This is a pragmatic choice. The formulation maximum is a regulatory ceiling rather than a maintenance reference, but the WHO parenteral-depot DDD, a utilisation unit, behaves poorly across depots with differing release kinetics. Reference doses for every agent, the reconciliation table, the injectable maxima and the WHO parenteral-depot DDDs are given in the Supplementary Material, each with its source.

### Phase 2: validation

#### Phase 2a: Clozapine component

We evaluated the clozapine term against measured plasma clozapine in the Clinical and Research Database (CRD) for Persistent Schizophrenia, an anonymised register of routine clinical and therapeutic-drug-monitoring data from the clozapine clinic at Cambridgeshire and Peterborough NHS Foundation Trust, which serves about 890,000 people (www.schizophrenias.group/database).

The authors assert that all procedures contributing to this work comply with the ethical standards of the relevant national and institutional committees on human experimentation and with the Helsinki Declaration of 1975, as revised in 2013. The database and its use for research were approved by the East of England Research Ethics Committee (references 13/EE/0121, 18/EE/0239 and 24/EE/0217). The database contains de-identified routine clinical data, and under these approvals individual consent was not required.

Repeated records of the same assay were removed. Analyses were restricted to a quality-controlled sample: prescribed dose ≥ 75 mg/day, plasma clozapine ≥ 0.10 mg/L, recorded sex and cigarettes/day, an available norclozapine level, a clozapine:norclozapine ratio between 0.5 and 3.0, agreement between the assay-linked and current dose (within 25% or 50 mg), and a concentration-to-dose ratio within 3 SD of the mean for the relevant sex–smoking stratum.

Because patients contributed repeated assessments, the primary analysis used linear mixed-effects models with a random intercept for patient. Steady-state concentration is proportional to dose divided by clearance, and the individualised target dose is itself proportional to clearance, being the dose that delivers the target concentration; the ratio of prescribed dose to target dose is therefore a clearance-adjusted dose. Both outcome and predictor were log-transformed accordingly; we verified that this specification fitted appreciably better than an untransformed predictor and adopted it throughout. Fit was summarised with marginal R² (variance explained by the fixed predictor) and conditional R² (fixed plus patient effects) following Nakagawa and Schielzeth,^25^ with Spearman correlation and discrimination (AUC) at 350 and 600 ng/mL as secondary measures. The two target schemes are not nested but have the same number of parameters, so they were compared directly by Akaike’s Information Criterion (AIC) and by a patient-level bootstrap.

Because the quality-control rule excludes concentration-to-dose outliers within sex–smoking strata, and the index is itself stratified by sex and smoking, we repeated the analysis without that exclusion, under minimal quality control, and using one randomly ordered observation per patient. We also asked what the index adds beyond its own ingredients by comparing it with models entering dose, sex and smoking as separate terms. Finally, because counting an antidepressant as polypharmacy is not meaningful for an antipsychotic index, assessments were split by the presence of a second antipsychotic rather than of any psychotropic. No patient was treated with fluvoxamine, an antidepressant that strongly inhibits CYP1A2.

#### Phase 2b: calibration of the non-clozapine reference

No comparable biomarker is available for the non-clozapine antipsychotics, so the reference can only be calibrated against other estimates of standard effective dosing. The systems under test are built from the licensed maximum and the WHO DDD; the criteria against which they are tested are derived from neither. The Gardner et al^2^ international consensus doses come from a survey of expert clinicians, and the near-maximal effective doses (ED95) of Leucht et al^14^ from dose-response curves fitted to randomised acute-phase trials.

For each candidate denominator system — %BNFmax, DDD, uncapped geometric mean, capped geometric mean, and a flat 1.5 × DDD — we computed the ratio of reference dose to criterion dose for every drug, and summarised each system by the standard deviation of the logarithm of that ratio across all drugs within that system. A systematic offset is unimportant on a ratio scale, since it rescales every score by the same constant; what matters is that the reference stands in a consistent proportion to the criterion from drug to drug, not that it is close to it in absolute terms. We then swept the cap from 1.0 to 2.5 times the DDD. This is calibration against other authors’ reference doses, not validation against patient outcomes.

## Results

### Clozapine validation cohort

The quality-controlled sample comprised 1,316 assessments from 255 patients, of which 765 were on clozapine without another antipsychotic and 551 with one (**Table 2**). Adjunct antipsychotics were present in 41.9% of assessments, antidepressants in 46.5% and mood stabilisers in 13.2%. Ethnicity, body weight and duration of clozapine treatment are not recorded in the extract.

**Table 2.** Characteristics of the validation cohort. Values are median (interquartile range) unless stated.

|  | **Value** |
|---|---|
| Assessments / patients | 1,316 / 255 |
| Assessments per patient | 5 (2–7) |
| Age at assay, years | 48.9 (40.7–56.5) |
| Male, % | 79.0 |
| Current smoker, % | 38.4 |
| Cigarettes/day among smokers | 20 (10–20) |
| Clozapine dose, mg/day | 300 (225–400) |
| Plasma clozapine, mg/L | 0.37 (0.25–0.52) |
| Plasma norclozapine, mg/L | 0.24 (0.16–0.33) |
| Clozapine:norclozapine ratio | 1.56 (1.25–1.92) |
| Adjunct antipsychotic, % | 41.9 |
| Antidepressant, % | 46.5 |
| Mood stabiliser, % | 13.2 |

### Individualised clozapine reference and plasma clozapine

The individualised reference explained substantially more of the variation in plasma clozapine than any conventional measure, and the two candidate schemes separated clearly (**Table 3**, **Figure 1**). Because chlorpromazine equivalents, olanzapine equivalents, the DDD and flat %BNFmax are all rescalings of clozapine dose, they rank patients identically and give one common fit, with a marginal R² of 0.166.

**Figure 1.**
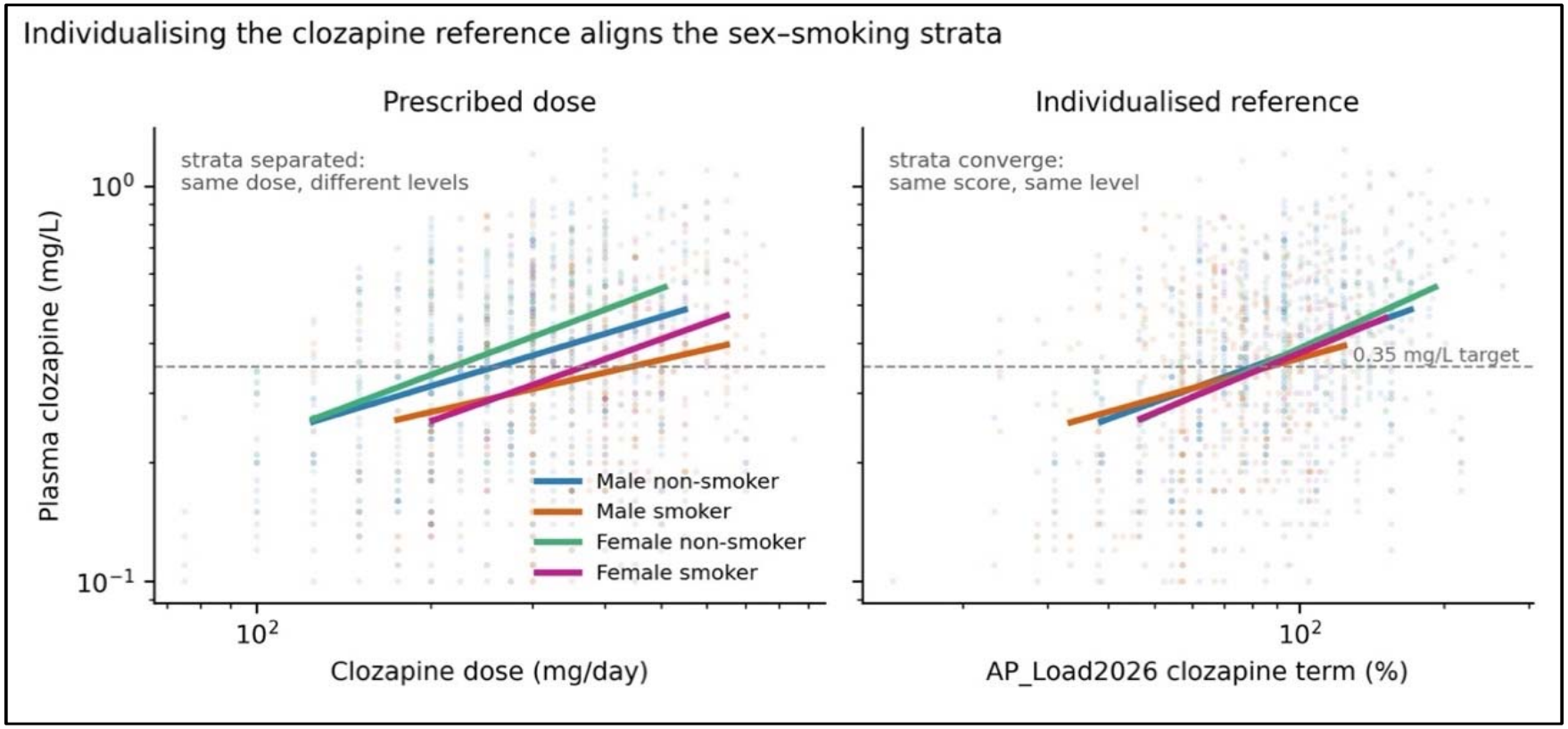
Plasma clozapine against prescribed dose (left) and against the individualised AP_Load2026 clozapine term (right), with fitted lines by sex–smoking stratum. Referencing the dose to an individualised target brings the four strata into alignment. Points are individual assessments; both axes are logarithmic.

**Table 3.**
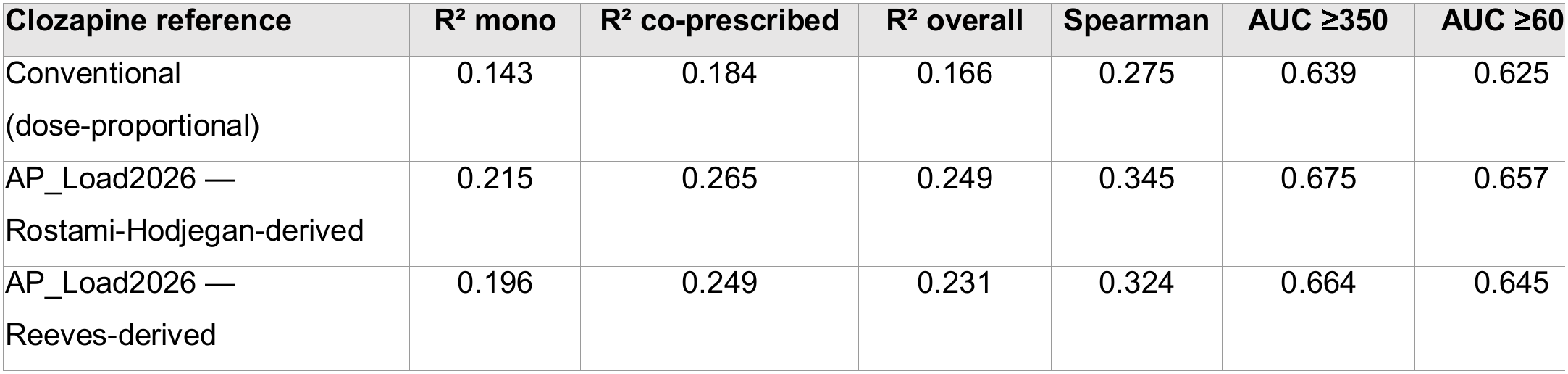
Validation of the AP_Load2026 clozapine term against plasma clozapine (mixed-effects models with a patient random intercept; log outcome and log predictor). The four conventional measures are rescalings of clozapine dose and give one common fit. Mono (monotherapy) = clozapine without another antipsychotic (n = 765); co-prescribed = clozapine with another antipsychotic (n = 551); overall n = 1,316.

| **Clozapine reference** | **R² mono** | **R² co-prescribed** | **R² overall** | **Spearman** | **AUC ≥350** | **AUC ≥60** |
|---|---|---|---|---|---|---|
| Conventional (dose-proportional) | 0.143 | 0.184 | 0.166 | 0.275 | 0.639 | 0.625 |
| AP_Load2026 — Rostami-Hodjegan-derived | 0.215 | 0.265 | 0.249 | 0.345 | 0.675 | 0.657 |
| AP_Load2026 — Reeves-derived | 0.196 | 0.249 | 0.231 | 0.324 | 0.664 | 0.645 |

The Rostami-Hodjegan-derived scheme raised this to 0.249 against 0.231 for the Reeves-derived scheme, with the same ordering in monotherapy, in antipsychotic co-prescription and across the secondary measures; conditional R² rose from 0.631 for the dose-proportional measures to 0.674 for the Rostami-Hodjegan-derived scheme. In formal comparison the Rostami-Hodjegan-derived scheme fitted better (ΔAIC 23.1; log-likelihood difference 11.6). In 1,000 patient-level bootstrap resamples it had the lower AIC in 99.1%, but the margin over the Reeves-derived scheme was modest and its confidence interval approached zero (ΔR² 0.018, 95% CI 0.001 to 0.034); the gain over dose-proportional measures was larger (ΔR² 0.082, 95% CI 0.044 to 0.120).

### Effect of sex and smoking on the clozapine term

Because the clozapine reference is a function of sex and smoking, and plasma clozapine is itself driven by sex and smoking, we asked whether the index achieves anything beyond entering those variables directly as covariates. Entering dose alone gave a marginal R² of 0.166; adding sex raised it to 0.176 and adding smoking to 0.249. The index alone gave the same 0.249, with two fewer fixed-effect terms (AIC 1224.7 versus 1226.2). Relative to male non-smokers, the dose required for a given concentration was roughly 1.4 to 1.6 times higher in smokers, whereas the female effect was small. A target fitted directly to this cohort’s own strata performed no better than the published anchors.

### Sensitivity analyses

Removing the sex–smoking-stratified concentration-to-dose exclusion left 1,339 assessments and gave a marginal R² of 0.150 for dose and 0.227 for the Rostami-Hodjegan-derived reference (ΔAIC 21.6 against the Reeves-derived scheme). Under minimal quality control (1,419 assessments) the corresponding marginal R² values were 0.141 and 0.211 (ΔAIC 21.6). Restricting to one observation per patient (n = 255) reduced all estimates but preserved the ordering (0.079, 0.136; ΔAIC 6.4). Absolute values were highest in the quality-controlled sample.

### Non-clozapine antipsychotics Behaviour across illustrative regimens

**Figure 2** shows how the three references treat the same everyday dose. Olanzapine 20 mg reads as 100% of the BNF maximum, 200% of the DDD, or 141% of the AP_Load2026 reference; risperidone 6 mg reads as 38%, 120% or 80%. The DDD over-weights ordinary doses, while %BNFmax understates precisely those agents whose licensed ceiling is far above real use. The comparison is extended to complete regimens, including clozapine augmentation and long-acting injectables, in Table 5 below.

**Figure 2.**
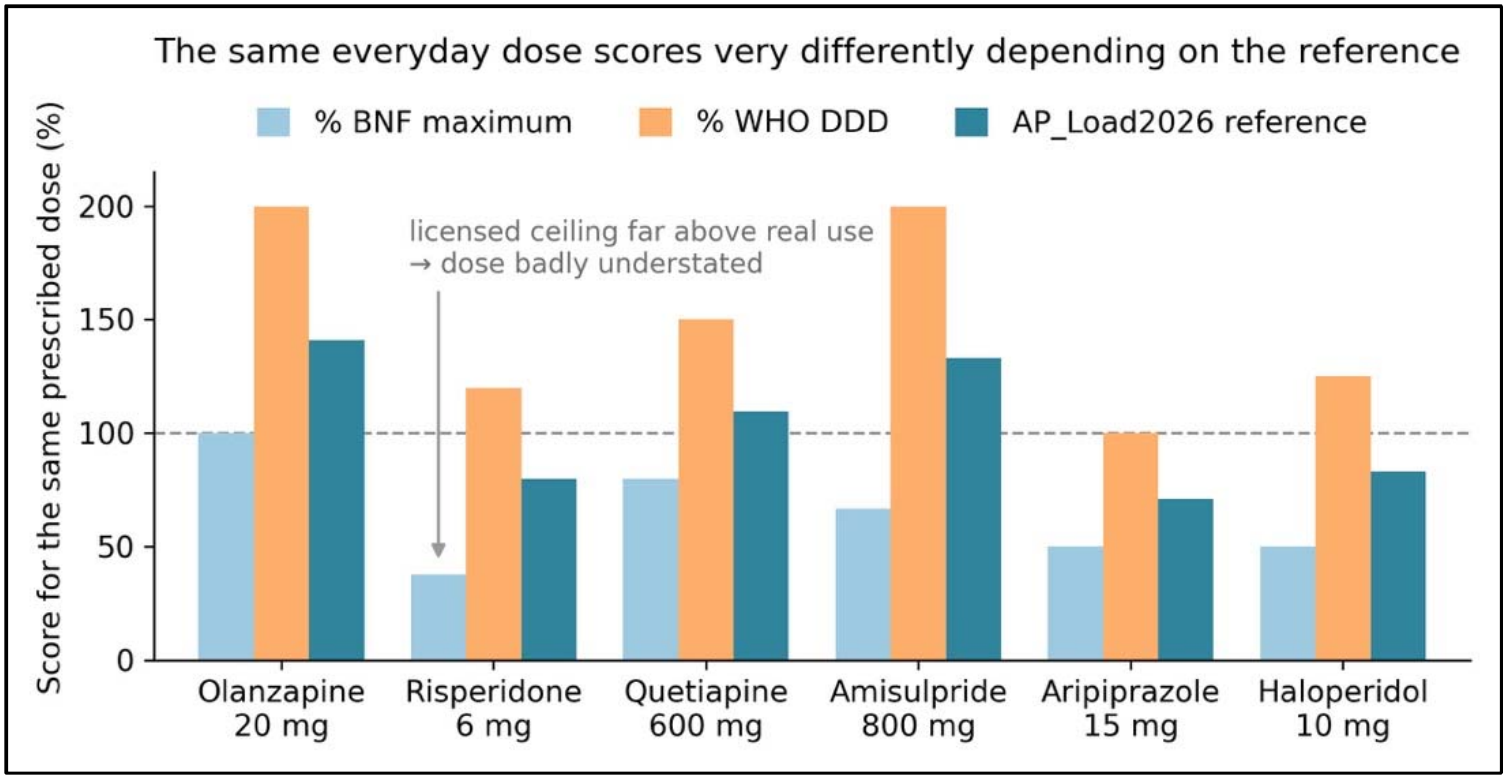
The same prescribed dose scored against three references. Values above 100% indicate a dose exceeding the reference. The licensed maximum understates agents such as risperidone whose ceiling sits far above the doses actually used, whereas the DDD over-weights ordinary doses. Only non-clozapine agents are shown, since the clozapine reference is individualised; clozapine is compared across the four sex–smoking strata in **Table 5**.

### Calibration of the non-clozapine reference

Against the Gardner consensus doses, covering fourteen agents, the capped geometric mean was the most consistent of the five candidate systems (SD of the log ratio 0.240), ahead of a flat 1.5 × DDD and the DDD itself (both 0.259) and the uncapped geometric mean (0.289); %BNFmax was clearly the least consistent (0.435) (**Table 4**). Against the near-maximal effective doses of Leucht et al,^14^ available for six agents, the four DDD-based systems were indistinguishable from one another (0.287 to 0.293) and %BNFmax was again the worst (0.337). Between 1.3 and 1.5 the cap had little influence, and above 1.5 consistency deteriorated towards that of the uncapped geometric mean. Against the Gardner criterion the optimum is shallow, with a minimum of 0.228 near 1.4 against 0.240 at the adopted value of 1.5; the Leucht criterion showed no discernible optimum (Figure 3). We retained the pre-specified value of 1.5 rather than the nominal minimum, since selecting a constant from the same data used to assess it would not be an independent choice.

**Figure 3.**
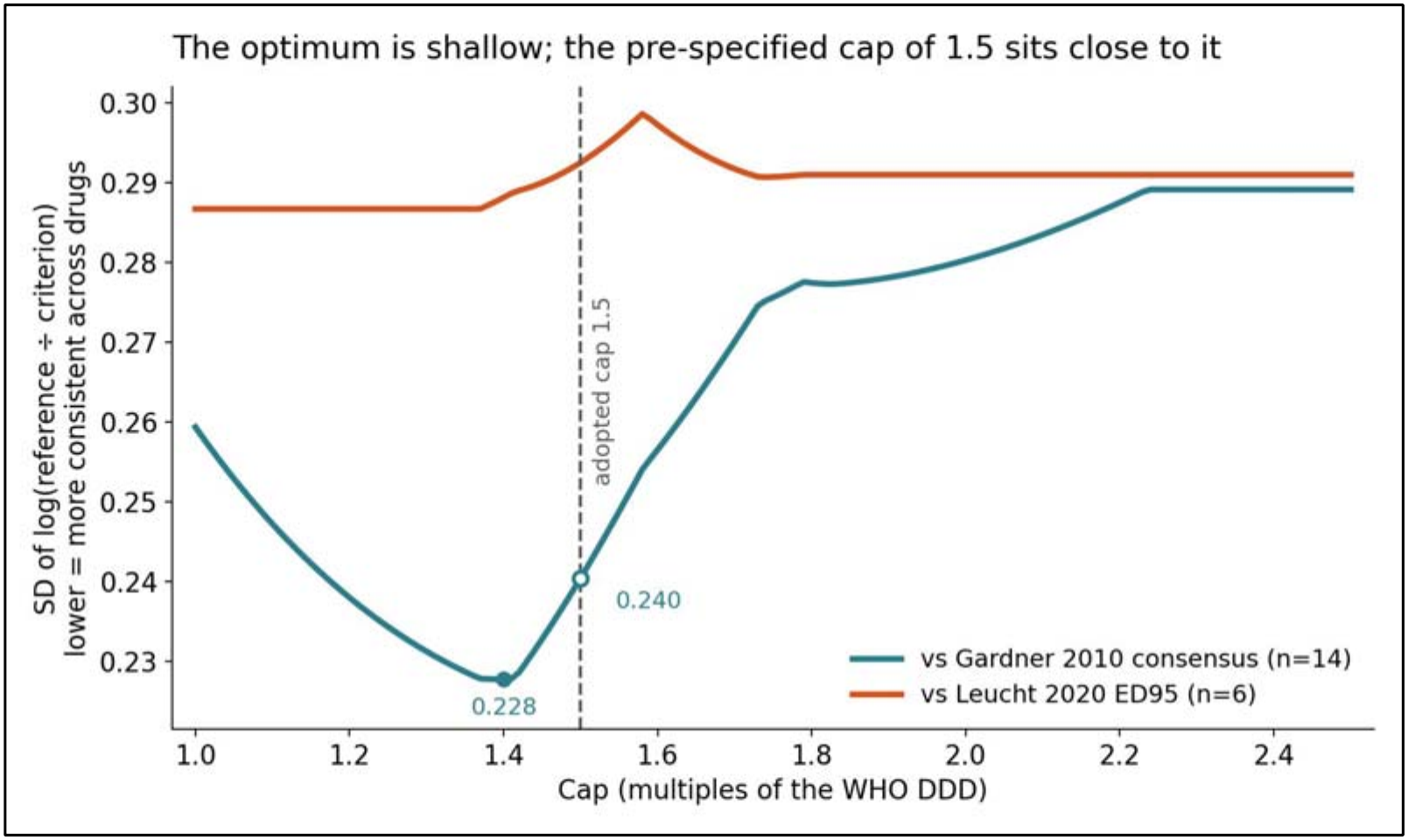
Sensitivity of the non-clozapine reference to the cap value. The optimum against the Gardner criterion is shallow: the minimum of 0.228 near 1.4 differs little from the 0.240 obtained at the adopted value of 1.5, and the difference is small relative to sampling variation across fourteen drugs. The Leucht criterion shows no discernible optimum.

**Table 4.**
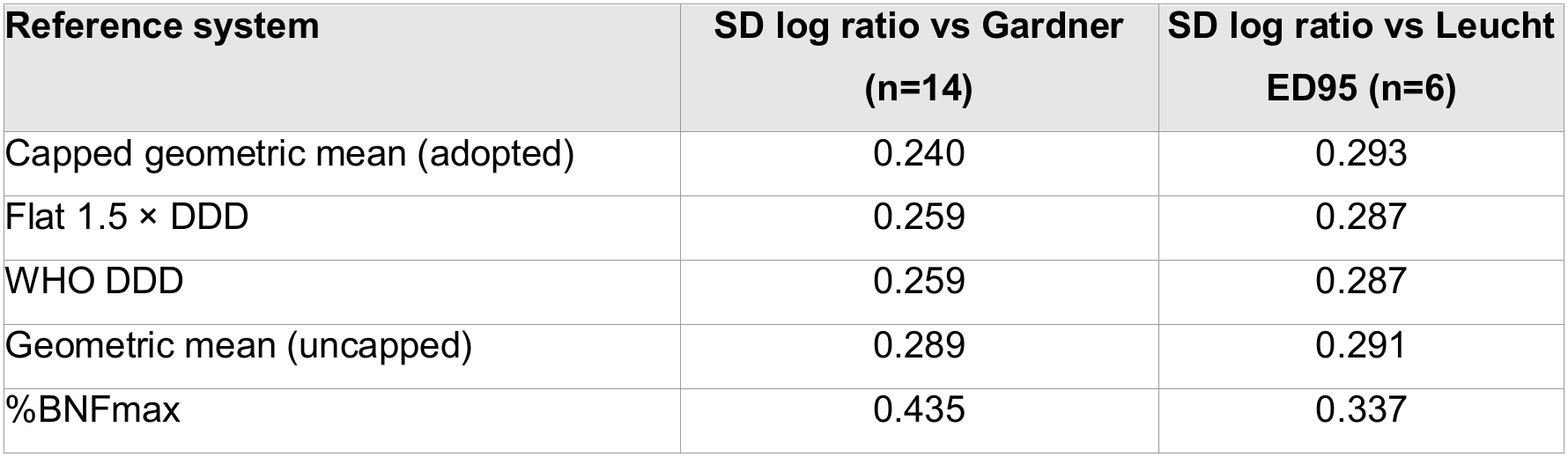
Consistency of five candidate reference-dose systems with published reference doses derived independently of both the DDD and the licensed maximum. Lower SD of the log ratio indicates a reference that is more consistently proportional to the external criterion across drugs; a uniform offset is unimportant on a ratio scale. The WHO DDD and the flat 1.5 × DDD give identical values by construction, since one is a constant multiple of the other, and both are shown for completeness.

### Behaviour of the index across complete regimens

Table 5 applies the final composite index to sixteen illustrative regimens: six non-clozapine monotherapies in a male non-smoker, the same clozapine dose across the four sex–smoking strata, and six clozapine augmentation regimens in a male smoker, two of them including a long-acting injectable.

**Table 5.**
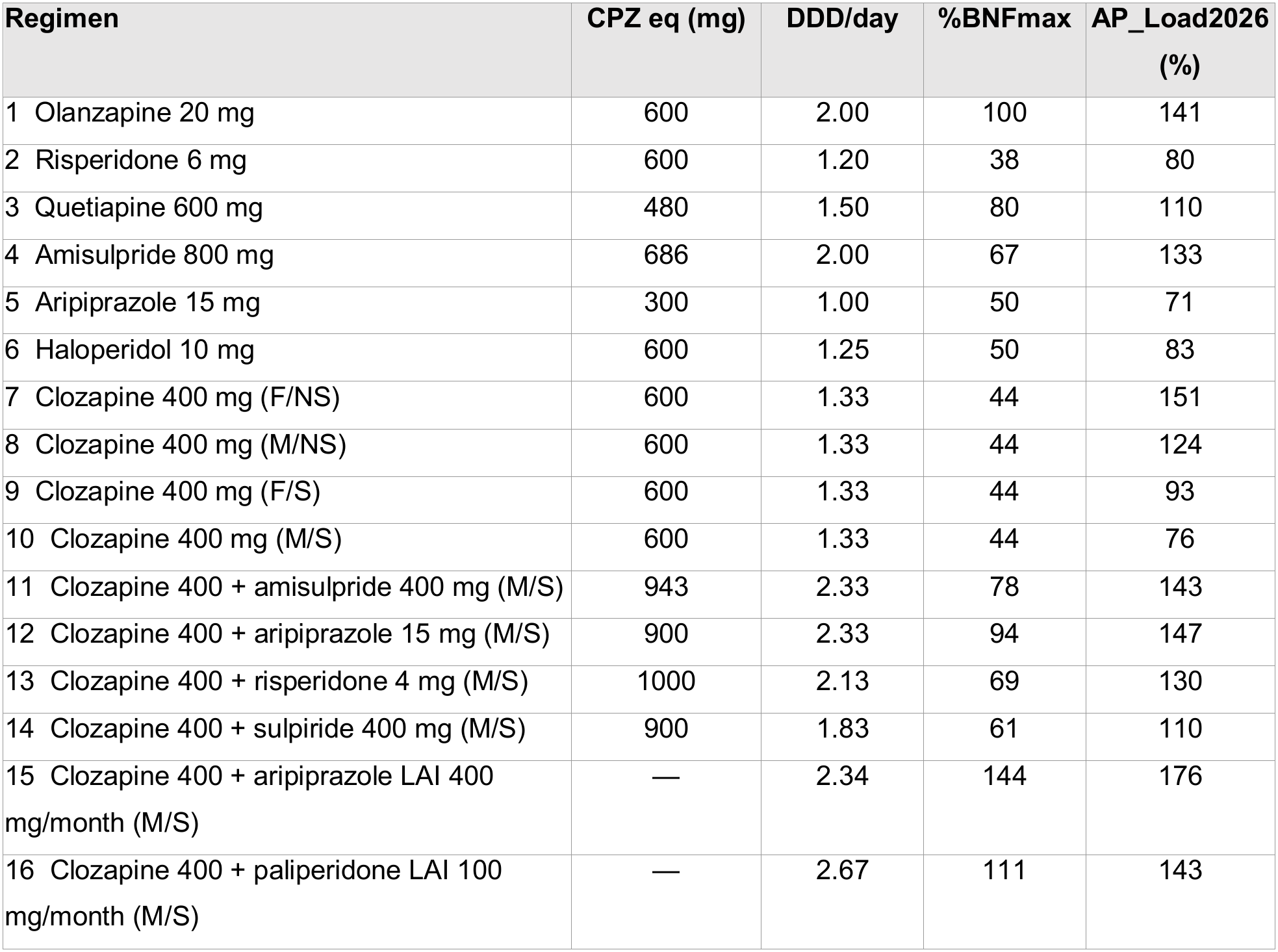
AP_Load2026 across sixteen illustrative regimens. Cases 1–6 are non-clozapine monotherapy in a male non-smoker; cases 7–10 are clozapine 400 mg/day in the four sex–smoking strata (F = female, M = male, NS = non-smoker, S = smoker at full induction); cases 11–16 are clozapine augmentation in a male smoker, cases 15 and 16 with a long-acting injectable (LAI). Chlorpromazine equivalents (CPZ eq) are taken from Gardner et al.^2^ %BNFmax scores clozapine against a flat 900 mg/day. Scores use the standard configuration: the Rostami-Hodjegan-derived clozapine anchors with the capped geometric-mean reference for other agents. AP_Load2026 scores are percentages of the reference dose, summed across agents without an upper cap. Values are illustrative, not empirical findings.

| **Regimen** | **CPZ eq (mg)** | **DDD/day** | **%BNFmax** | **AP_Load2026 (%)** |
|---|---|---|---|---|
| 1 Olanzapine 20 mg | 600 | 2.00 | 100 | 141 |
| 2 Risperidone 6 mg | 600 | 1.20 | 38 | 80 |
| 3 Quetiapine 600 mg | 480 | 1.50 | 80 | 110 |
| 4 Amisulpride 800 mg | 686 | 2.00 | 67 | 133 |
| 5 Aripiprazole 15 mg | 300 | 1.00 | 50 | 71 |
| 6 Haloperidol 10 mg | 600 | 1.25 | 50 | 83 |
| 7 Clozapine 400 mg (F/NS) | 600 | 1.33 | 44 | 151 |
| 8 Clozapine 400 mg (M/NS) | 600 | 1.33 | 44 | 124 |
| 9 Clozapine 400 mg (F/S) | 600 | 1.33 | 44 | 93 |
| 10 Clozapine 400 mg (M/S) | 600 | 1.33 | 44 | 76 |
| 11 Clozapine 400 + amisulpride 400 mg (M/S) | 943 | 2.33 | 78 | 143 |
| 12 Clozapine 400 + aripiprazole 15 mg (M/S) | 900 | 2.33 | 94 | 147 |
| 13 Clozapine 400 + risperidone 4 mg (M/S) | 1000 | 2.13 | 69 | 130 |
| 14 Clozapine 400 + sulpiride 400 mg (M/S) | 900 | 1.83 | 61 | 110 |
| 15 Clozapine 400 + aripiprazole LAI 400 mg/month (M/S) | — | 2.34 | 144 | 176 |
| 16 Clozapine 400 + paliperidone LAI 100 mg/month (M/S) | — | 2.67 | 111 | 143 |

Reference implementations of AP_Load2026 in Python, R and SPSS, together with the full reference tables and the sixteen regimens of Table 5 as a worked example, are openly available (see Data availability).

## Discussion

AP_Load2026 expresses each prescribed dose as a percentage of a documented, dated reference dose and sums those percentages uncapped; its two components were constructed and evidenced differently by design.

For clozapine, whose effective dose is set by clearance rather than by a fixed milligram ceiling, referencing the dose to a target individualised for sex and cigarettes per day tracked measured plasma clozapine considerably better than any dose-proportional measure (marginal R² 0.249 versus 0.166; ΔR² 0.082, 95% CI 0.044 to 0.120). The advantage held under every quality-control variant and under co-prescription. Of the two published sets, the Rostami-Hodjegan-derived scheme fitted better than the Reeves-derived, consistently in direction though modestly in size.

For the other antipsychotics, where no biomarker can arbitrate, the capped geometric mean of the DDD and the licensed maximum was the most consistent of five candidate references against the Gardner consensus doses and indistinguishable from the other DDD-based systems against the Leucht ED95 criterion; %BNFmax was the least consistent against both. Across sixteen illustrative regimens the index separated prescriptions that conventional measures score identically, most markedly under clozapine augmentation.

### Comparison with existing measures and evidence

The smoking effect observed here, a dose requirement roughly 1.4 to 1.6 times higher in smokers, lies within the published pharmacokinetic range,^22,26^ whereas the female effect was small. The Rostami-Hodjegan-derived smoking effect of about +62% matches this closely, while the Reeves-derived +30% under-corrects and separates the key strata less well. The small female effect is consistent with the view that much of the apparent sex difference in clozapine clearance is carried by smoking prevalence and body size rather than by sex itself.^17^ That a target fitted to this cohort’s own strata performed no better than the published anchors suggests that the external coefficients are already close to the achievable ceiling here.

The share of variance explained is still modest in absolute terms, as plasma clozapine is also shaped by adherence, sampling interval, assay variation, body weight, age, inflammation, caffeine and CYP1A2 genotype, none of which a prescription-based index can fully capture. The informative comparison is therefore relative, against dose-proportional measures such as the DDD or %BNFmax, rather than absolute. Nor is the index a repackaging of its own ingredients: it reproduced a model entering dose, sex and smoking as separate terms with two fewer fixed-effect terms. Its advantage is that clozapine dose is normalised for two major determinants of its pharmacokinetics. Without such normalisation, a fixed-milligram measure differentially misclassifies clozapine exposure according to sex and smoking, potentially confounding analyses in clozapine-enriched samples, particularly when sex or smoking are themselves variables of interest (e.g. smoking in studies of cognition).

Indeed, normalising a raw measurement against a demographically referenced expectation is already routine elsewhere in schizophrenia research but has not been applied to dose. Cognition is the closest parallel. The Brief Assessment of Cognition in Schizophrenia (BACS) is reported as a composite z-score standardised against age- and sex-matched normative data.^27,28^ The normalisation exists so that the same raw performance carries the same meaning in different people. AP_Load2026 does the same for prescribed dose, referencing 400 mg of clozapine to what that dose means in a person of that sex and smoking status, and placing the remaining antipsychotics on the same percentage scale through an explicit, if conventional, reference rule.

### What the index does and does not represent

AP_Load2026 is a prescribed-dose normalisation framework for research. It is not an equipotency scale, a receptor-occupancy model, or a means of predicting an individual’s plasma concentration; where a plasma level is available it remains preferable.^29^ It measures antipsychotic dose only; other dimensions of medication burden, such as anticholinergic load, require dedicated instruments.^30,31^ Nor is it a prescribing or regulatory instrument. UK high-dose thresholds rest on the same percentage-of-maximum arithmetic^32^ and they remain anchored to the licensed maximum. Whether a normalised load could serve that purpose is a separate question, which we do not address here.

A score of 100 denotes one reference-dose unit, anchored to maintenance rather than acute dosing because that is what most clinical studies sample.^33^ It is not a therapeutic threshold, and its correspondence with independent estimates of standard effective dosing is not uniform across drugs: against the Gardner consensus doses the reference ranges from 0.71 of the criterion (aripiprazole, olanzapine, ziprasidone) to 1.50 (sulpiride), with a geometric mean of 0.90 (**Supplementary Table S3**). On the non-clozapine side the cap does most of the work: risperidone’s uncapped geometric mean of 8.9 mg is pulled upward by a 16 mg licensed ceiling, and capping returns it to 7.5 mg, within the range where utilisation, prescribing and efficacy converge,^9,14,34^ so a 6 mg dose scores 80% rather than 38%. **Supplementary Table S8** restates the reference for every agent as the prescribed dose, in milligrams, that corresponds to an AP_Load2026 of 100.

Summation is a convention too. Adding percentages assumes that two agents at half their reference doses are equivalent, as exposure, to one agent at its reference dose. Nothing here tests that, and the plasma-clozapine criterion cannot adjudicate it, since a co-prescribed drug does not alter clozapine concentrations. We adopt it because it is transparent and reproducible, and because potency- or occupancy-weighted alternatives reintroduce the contested ratios the index avoids. Since estimated D2 occupancy rises non-linearly with dose,^35^ a summed score likely over-states combined exposure at high totals.

Because sex and smoking sit inside the exposure definition, AP_Load2026 is not a pure treatment variable: two patients on identical clozapine doses score differently when one of them smokes, and since smoking may also affect the outcome directly, sex, smoking and other confounders must be modelled alongside it. Because the arithmetic is formulary-agnostic, only the reference table need be swapped outside the UK — a flexibility that becomes a hazard if one centre uses BNF maxima and another FDA labels. National maxima do differ materially: haloperidol is capped at 20 mg/day in the current BNF, whereas US labelling describes oral doses of up to 100 mg/day in severely resistant cases, and agents licensed across Europe, such as amisulpride and sulpiride, have no US oral licence at all.

We therefore distinguish a reference configuration, using the published table in the Supplementary Material and intended for cross-study comparison, from a locally recalibrated configuration, which should be labelled as such; the suffix 2026 marks the table intentionally. The index is intended for investigators treating antipsychotic medication as a covariate, a sample descriptor or an exposure of interest: studies of negative symptoms, cognition and motivation,^36,37^ early-psychosis cohorts,^38^ register-based studies of physical-health outcomes,^10,39^ and clozapine-treated cohorts.

### Limitations

AP_Load2026 is a prescribed-dose proxy rather than a biological measure. The clozapine target uses sex and cigarettes per day as markers of CYP1A2 activity but does not capture genotype, age, weight, ancestry, caffeine intake, inflammation, adherence or interacting co-medications. The validation is narrow by design. It draws on a single specialist service, explains only a modest share of plasma variance and directly validates only the clozapine component. Its strength is the relatively large number of quality-controlled therapeutic drug monitoring assessments. The continuous smoking specification is barely exercised here: only about one smoking record in seven fell between one and nine cigarettes per day, so the interpolation operates almost entirely at its saturated value and the comparison is in practice binary. The non-clozapine reference was calibrated against other authors’ published reference doses rather than patient outcomes, so it establishes concordance and cannot show that the reference predicts efficacy or tolerability; the handling of long-acting injectables and the optional antidepressant, mood-stabiliser and benzodiazepine loads are not validated at all. Indices of this kind are better established on larger, multi-site datasets, and these figures are a first calibration, not a definitive one.

A related limitation concerns interpretation of AP_Load2026 as a proxy for clinical risk rather than dose exposure. Because the index sums normalised dose across concurrently prescribed antipsychotics, augmentation regimens will almost by construction score higher than monotherapy, regardless of whether the added agent contributes meaningful additional antipsychotic exposure. Aripiprazole, for instance, is commonly added to clozapine to mitigate sedation or compulsive behaviour rather than to strengthen antipsychotic effect,^40,41^ yet it contributes fully to the summed score, illustrating that a high index value can reflect adverse-effect burden as much as therapeutic coverage. AP_Load2026 has not been validated as a measure of clinical risk or treatment resistance, and augmentation status should be considered when interpreting or modelling the index.

## Conclusions

AP_Load2026 offers a transparent, reproducible and versioned way to normalise prescribed antipsychotic dose for research. Its principal advance is an individualised clozapine reference that tracks measured plasma clozapine considerably better than raw dose, the DDD or flat %BNFmax, carrying known pharmacokinetic variation in one interpretable variable with coefficients fixed in advance. The reference for other antipsychotics is a calibrated convention rather than a validated measure. It is a documented research proxy, complementary to therapeutic drug monitoring and genotyping where those are available, and not a clinical or regulatory instrument.

## Data Availability

All data produced in the present study are available upon reasonable request to the authors.

Reference implementations of AP_Load2026 in Python, R and SPSS, together with the complete dated reference tables, the sixteen worked regimens of Table 5 and the tests that reproduce them, are openly available under version control at https://github.com/eegea73/AP_load and www.schizophrenias.group/ap_load. The Clinical and Research Database extract underlying the clozapine validation consists of routinely collected clinical data and cannot be shared publicly; enquiries should be directed to the corresponding author.

## Acknowledgments

Claude (Anthropic) was used to assist with language editing, reference checking and drafting of the reference code implementations. All outputs were reviewed and verified by the authors, who take full responsibility for the content of this manuscript.

## Author Contributions

EFE, CJ and NW conceived and designed the study. CJ and EFE assembled and curated the clinical and therapeutic-drug-monitoring extract. EFE and NW performed the statistical analysis. EFE wrote the reference implementations. EFE wrote the first draft. All authors commented on and edited the manuscript, approved the final version, and agree to be accountable for all aspects of the work.

