## Supplementary Material for "The Antipsychotic Load (AP_Load2026): a normalised prescribed-dose index for schizophrenia research"

**Supplementary Material — AP_Load2026**

This appendix gives the reference dose used for every agent and its source (Tables S1, S2), the calibration of those references against external criteria (Table S3), the optional non-antipsychotic loads (Tables S4–S6), the statistical specification (Table S7) the prescribed dose corresponding to a score of 100 for every agent (Table S8), the consistency of five candidate reference systems with the external criteria (Table S9), the same prescribed doses scored against three references (Figure S1) and the sensitivity of the non-clozapine reference to the cap (Figure S2). The clozapine individualised target doses are given in Table 1 of the main text.

### 1. The standard configuration

AP_Load2026 scores each antipsychotic as (prescribed daily dose) ÷ (reference dose) × 100 and sums the percentages across agents without an upper cap. One configuration is recommended for uniform use:

1. **Clozapine.** Referenced to the individualised target dose for a plasma clozapine of 0.35 mg/L (350 ng/mL) using the Rostami-Hodjegan-derived anchors (Rostami-Hodjegan et al., 2004; main-text Table 1): female non-smoker 265, male non-smoker 323, female smoker 431, male smoker 525 mg/day. The clozapine score is not capped at 100.
2. **Smoking input.** Entered continuously as cigarettes per day, target(sex, cig) = T_NS + (T_S − T_NS) × min(1, cig/10), so induction saturates at ten cigarettes per day.
3. **Other oral antipsychotics.** Referenced to the geometric mean of the BNF (or national label) maximum and the WHO DDD, √(BNFmax × DDD), capped at 1.5 × DDD; agents with no assigned WHO DDD keep their label maximum (Table S1).
4. **Long-acting injectables.** Referenced to the licensed formulation maximum for the same dosing interval, so that the prescribed dose per interval is used directly without conversion to a daily equivalent (Table S2).
5. **Other classes.** Antidepressant, mood-stabiliser and benzodiazepine loads (Tables S4–S6) use the same arithmetic against the BNF maximum. They are descriptive adjuncts only: they are not validated and form no part of AP_Load2026.

Alternative configurations — the %BNFmax or DDD denominators, and the Reeves-derived clozapine anchors — can be reproduced from the reference doses tabulated below and the anchors in main-text Table 1; they are recorded for transparency, not offered as equivalent options.

### 2. The reconciliation rule for non-clozapine agents

The rule and its rationale are given in the main text; in brief, the geometric mean lies an equal proportional distance from each reference, so neither dominates by being the larger number, and it returns the BNF maximum where the two coincide. The cap binds wherever the BNF-to-DDD ratio exceeds 2.25, which is the point at which the geometric mean would otherwise pass 1.5 × DDD; these agents are marked in Table S1. The rule applies uniformly to every agent with a WHO DDD.

The UK BNF (2026) was the primary source, applied uniformly. The Summary of Product Characteristics (SmPC; eMC, or AEMPS/CIMA), FDA prescribing information or other national labels were used where a UK BNF value was unavailable.

### Table S1. Antipsychotic reference doses

| **Antipsychotic** | **BNF / label max (mg/day)** | **WHO DDD (mg/day)** | **Ratio** | **Geometric mean √(BNFmax×DDD)** | **AP_Load2026 reference (mg/day)** | **Basis** | **Source** |
| --- | --- | --- | --- | --- | --- | --- | --- |
| Amisulpride | 1200 | 400 | 3.00 | 693 | 600 | 1.5 × DDD (cap) | BNF |
| Aripiprazole (oral) | 30 | 15 | 2.00 | 21.2 | 21.2 | Geometric mean | BNF |
| Asenapine | 20 | 20 | 1.00 | 20 | 20 | Geometric mean (= label max) | FDA / SmPC |
| Blonanserin | 24 | — | — | — | 24 | Label max (no WHO DDD) | PMDA (Japan) |
| Brexpiprazole | 4 | — | — | — | 4 | Label max (no WHO DDD) | FDA |
| Cariprazine | 6 | — | — | — | 6 | Label max (no WHO DDD) | FDA / SmPC |
| Chlorpromazine | 1000 | 300 | 3.33 | 548 | 450 | 1.5 × DDD (cap) | BNF |
| Clozapine | 900 | 300 | 3.00 | — | Individualised | Target for 0.35 mg/L (main-text Table 1) | BNF |
| Flupentixol (oral) | 18 | 6 | 3.00 | 10.4 | 9.0 | 1.5 × DDD (cap) | BNF |
| Haloperidol (oral) | 20 | 8 | 2.50 | 12.6 | 12 | 1.5 × DDD (cap) | BNF |
| Iloperidone | 24 | — | — | — | 24 | Label max (no WHO DDD) | FDA |
| Lumateperone | 42 | — | — | — | 42 | Label max (no WHO DDD) | FDA |
| Lurasidone | 148 | 60 | 2.47 | 94 | 90 | 1.5 × DDD (cap) | BNF |
| Olanzapine | 20 | 10 | 2.00 | 14.1 | 14.1 | Geometric mean | BNF |
| Paliperidone (oral) | 12 | 6 | 2.00 | 8.5 | 8.5 | Geometric mean | BNF |
| Pimozide | 20 | 4 | 5.00 | 8.9 | 6.0 | 1.5 × DDD (cap) | BNF |
| Quetiapine | 750 | 400 | 1.88 | 548 | 548 | Geometric mean | BNF |
| Risperidone | 16 | 5 | 3.20 | 8.9 | 7.5 | 1.5 × DDD (cap) | BNF |
| Sulpiride | 2400 | 800 | 3.00 | 1386 | 1200 | 1.5 × DDD (cap) | BNF |
| Trifluoperazine | 20 | 20 | 1.00 | 20 | 20 | Geometric mean (= label max) | SmPC |
| Xanomeline–trospium | 250 | — | — | — | 250 | Label max (no WHO DDD); xanomeline component | FDA |
| Ziprasidone | 160 | 80 | 2.00 | 113 | 113 | Geometric mean | FDA / SmPC |
| Zuclopenthixol (oral) | 150 | 30 | 5.00 | 67 | 45 | 1.5 × DDD (cap) | BNF |
| **Agents outside the UK formulary** |  |  |  |  |  |  |  |
| Fluphenazine (oral) | — | 10 | — | — | 15 | 1.5 × DDD (oral form discontinued in the UK) | WHO |
| Levomepromazine | — | 300 | — | — | 450 | 1.5 × DDD (no current UK max) | WHO |
| Penfluridol | — | 6 | — | — | 9 | 1.5 × DDD (weekly oral; daily equivalent) | WHO |

*Ratio = BNF or label maximum ÷ WHO DDD; the cap is binding where it exceeds 2.25. The AP_Load2026 reference is the dose that scores 100: a prescribed dose is divided by it and multiplied by 100. Agents outside the UK formulary are those that may be met in non-UK datasets, having a WHO DDD but no current UK maximum, so no geometric mean can be formed; the reference defaults to the cap itself, 1.5 × DDD, the upper bound the rule would impose in any case, and a national label maximum can be substituted where one exists. Penfluridol is dosed weekly and its per-interval dose is converted to a daily equivalent before scoring. Xanomeline–trospium is scored on its xanomeline component; the peripherally restricted trospium component is not counted. Doses of this combination are conventionally quoted as the pair (for example 125 mg/30 mg twice daily), so the value tabulated is the daily xanomeline total. Agents without an assigned WHO DDD would gain reconciliation automatically if a WHO or local DDD were substituted.*

### Table S2. Long-acting injectables and depots

| **Drug** | **Interval** | **AP_Load2026 reference (maximum per interval)** | **WHO depot DDD (mg/day)** | **Source** |
| --- | --- | --- | --- | --- |
| Aripiprazole | monthly | 400 mg | 13.3 | BNF; WHO |
| Aripiprazole | 2-monthly | 960 mg | 13.3 | BNF; WHO |
| Paliperidone* | monthly | 150 mg | 2.5 | BNF; WHO |
| Paliperidone* | 3-monthly | 525 mg | 2.5 | BNF; WHO |
| Paliperidone* | 6-monthly | 1000 mg | 2.5 | SmPC (Byannli) |
| Risperidone | 2-weekly | 50 mg | 2.7 | BNF; WHO |
| Olanzapine | 2-weekly | 300 mg | — | SmPC |
| Olanzapine | 4-weekly | 405 mg | — | SmPC |
| Haloperidol decanoate | 4-weekly | 300 mg | 3.3 | BNF; WHO |
| Flupentixol decanoate | weekly | 400 mg | — | BNF |
| Zuclopenthixol decanoate | weekly | 600 mg | — | BNF |
| Fluphenazine decanoate | 2-weekly | 100 mg | — | BNF / SmPC |

*The AP_Load2026 reference is the licensed formulation maximum for the same dosing interval, tabulated in the third column: a dose per interval is divided by it and multiplied by 100. WHO parenteral-depot DDDs (mg/day — the average recommended dose divided by the dosing interval) are shown for reference only; a DDD-based depot score would divide the per-interval dose, converted to mg/day, by the depot DDD. "—" indicates no depot DDD retrieved. * Paliperidone doses are expressed as paliperidone (active moiety), not paliperidone palmitate: the 6-monthly maximum of 1000 mg paliperidone corresponds to 1560 mg paliperidone palmitate.*

### Table S3. Calibration of the reference dose against external criteria

| **Drug** | **AP_Load2026 reference (mg/day)** | **Gardner consensus dose** | **Ratio (Gardner)** | **Leucht ED95** | **Ratio (Leucht)** |
| --- | --- | --- | --- | --- | --- |
| Amisulpride | 600 | 700 | 0.86 | 537 | 1.12 |
| Aripiprazole | 21.2 | 30 | 0.71 | 11.5 | 1.84 |
| Chlorpromazine | 450 | 600 | 0.75 | — | — |
| Flupentixol | 9.0 | 10 | 0.90 | — | — |
| Haloperidol | 12.0 | 10 | 1.20 | 6.3 | 1.90 |
| Olanzapine | 14.1 | 20 | 0.71 | 15.2 | 0.93 |
| Paliperidone* | 8.5 | 9 | 0.94 | — | — |
| Pimozide | 6.0 | 8 | 0.75 | — | — |
| Quetiapine | 547.7 | 750 | 0.73 | 482 | 1.14 |
| Risperidone | 7.5 | 6 | 1.25 | 6.26 | 1.20 |
| Sulpiride | 1200 | 800 | 1.50 | — | — |
| Trifluoperazine | 20.0 | 20 | 1.00 | — | — |
| Ziprasidone | 113.1 | 160 | 0.71 | — | — |
| Zuclopenthixol | 45.0 | 50 | 0.90 | — | — |

*Criteria are the Gardner et al. (2010) international consensus dose and the near-maximal effective dose (ED95) of Leucht et al. (2020), both derived independently of the WHO DDD and of the licensed maximum. Ratio = AP_Load2026 reference ÷ criterion. Across the fourteen agents the adopted reference sits at a geometric-mean ratio of 0.90 to the Gardner consensus dose and 1.31 to the acute ED95. The consistency of the five candidate reference systems against these criteria is given in Table S9; note there that the WHO DDD and a flat 1.5 × DDD necessarily share an identical SD of the log ratio, because multiplying every reference by a constant shifts the mean of the log ratios but cannot change their dispersion — they differ in centring, not in consistency.*

### Optional non-antipsychotic loads (descriptive adjuncts)

*These three loads use the same arithmetic — prescribed daily dose as a percentage of the BNF maximum, summed — but were not validated in this work and form no part of AP_Load2026. They are provided as optional exploratory outputs.*

Table S4. Antidepressants

| **Drug** | **BNF max (mg/day)** | **Source** |
| --- | --- | --- |
| Amitriptyline | 200 | BNF |
| Bupropion | 300 | SmPC (European) |
| Citalopram | 40 | BNF |
| Clomipramine | 250 | BNF |
| Duloxetine | 120 | BNF |
| Escitalopram | 20 | BNF |
| Fluoxetine | 60 | BNF |
| Fluvoxamine | 300 | BNF |
| Mirtazapine | 45 | BNF |
| Paroxetine | 50 | BNF |
| Sertraline | 200 | BNF |
| Trazodone | 600 | BNF |
| Venlafaxine | 375 | BNF |
| Vortioxetine | 20 | BNF |

#### Table S5. Mood stabilisers

| **Drug** | **Maximum (mg/day)** | **Source / note** |
| --- | --- | --- |
| Carbamazepine | 1600 | BNF (bipolar) |
| Lamotrigine | 200 | BNF (bipolar monotherapy) |
| Lithium | 2000 | BNF (nominal; salt-dependent — weak proxy) |
| Oxcarbazepine | 2400 | BNF (off-label as mood stabiliser) |
| Valproate | 2000 | BNF (bipolar maintenance) |

*Lithium is titrated to serum concentration and milligram doses are not comparable across salts and brands; we recommend excluding it from a default mood-stabiliser load, or flagging it explicitly.*

#### Table S6. Benzodiazepines (standing doses only)

| **Drug** | **BNF / SmPC max (mg/day)** | **Source / note** |
| --- | --- | --- |
| Alprazolam | 4 | SmPC (European) |
| Bromazepam | 36 | SmPC (European) |
| Clobazam | 60 | BNF (epilepsy) |
| Clonazepam | 8 | BNF (epilepsy maximum) |
| Diazepam | 30 | BNF |
| Lorazepam | 4 | BNF |
| Nitrazepam | 10 | BNF |
| Oxazepam | 120 | BNF |
| Temazepam | 40 | BNF |

*As-required (PRN) entries are ignored; only standing doses contribute. Benzodiazepine maxima are frequently indication-dependent — the clonazepam maximum reflects epilepsy, for example — so this domain is coarser than AP_Load.*

### Table S7. Statistical specification

Given in full so that the analyses can be reproduced exactly.

| **Element** | **Specification** |
| --- | --- |
| Clozapine target function | target(sex, cig) = T_NS + (T_S − T_NS) × min(1, cig/10), where T_NS and T_S are the non-smoker and smoker anchors for that sex (main-text Table 1) and cig is cigarettes per day. |
| Clozapine term | AP_Load clozapine score = prescribed dose ÷ target(sex, cig) × 100. |
| Primary quality control | Applied in order: dose ≥ 75 mg/day; plasma clozapine ≥ 0.10 mg/L; norclozapine present; clozapine:norclozapine ratio 0.5–3.0; \|assay-linked dose − current treatment dose\| ≤ max(25% of treatment dose, 50 mg), whichever tolerance is larger; concentration-to-dose ratio within 3 SD of the mean of its sex–smoking stratum. Repeated records of the same assay (same patient, date, dose and results) were removed first. |
| Minimal quality control | Dose ≥ 75 mg/day and plasma clozapine ≥ 0.10 mg/L only; no ratio, dose-agreement or stratum-outlier criteria. |
| Primary model | log(plasma clozapine) ~ log(predictor) + (1 \| patient), linear mixed-effects, maximum likelihood. The log-log form follows from the conceptual model that concentration is proportional to dose divided by target dose, and fitted appreciably better than an untransformed predictor (ΔAIC 39.6). |
| Comparator model | log(plasma clozapine) ~ log(dose) + sex + smoking + (1 \| patient), where smoking is binary current-smoker status. A variant using cigarettes/day as a continuous term is also reported. |
| Marginal and conditional R² | Marginal R² = var(fixed-effect fitted values) ÷ [var(fixed-effect fitted values) + random-intercept variance + residual variance]; conditional R² adds the random-intercept variance to the numerator (Nakagawa and Schielzeth, 2013). A single definition is used throughout. |
| Model comparison | Akaike Information Criterion on maximum-likelihood fits. The two clozapine schemes are not nested but carry the same number of parameters, so AIC differences reduce to twice the log-likelihood difference. |
| Bootstrap | 1,000 resamples of patients (not observations) with replacement, preserving each patient’s full set of assessments; percentile 95% confidence intervals. |
| Denominator calibration | For each candidate system and drug, deviation = log(reference dose ÷ criterion dose). Systems compared by the SD of that deviation across drugs (consistency); the geometric mean of the ratio is reported separately (centring). Cap swept from 1.0 to 2.5 × DDD. |

### Table S8. Prescribed dose corresponding to an AP_Load2026 of 50, 100 and 150

| **Agent** | **AP_Load2026 = 50** | **AP_Load2026 = 100 (reference dose)** | **AP_Load2026 = 150** |
| --- | --- | --- | --- |
| **Oral antipsychotics (mg/day)** | | | |
| Amisulpride | 300 | 600 | 900 |
| Aripiprazole | 10.6 | 21.2 | 31.8 |
| Asenapine | 10 | 20 | 30 |
| Blonanserin | 12 | 24 | 36 |
| Brexpiprazole | 2 | 4 | 6 |
| Cariprazine | 3 | 6 | 9 |
| Chlorpromazine | 225 | 450 | 675 |
| Flupentixol | 4.5 | 9.0 | 13.5 |
| Fluphenazine † | 7.5 | 15 | 22.5 |
| Haloperidol | 6 | 12 | 18 |
| Iloperidone | 12 | 24 | 36 |
| Levomepromazine † | 225 | 450 | 675 |
| Lumateperone | 21 | 42 | 63 |
| Lurasidone | 45 | 90 | 135 |
| Olanzapine | 7.1 | 14.1 | 21.2 |
| Paliperidone | 4.2 | 8.5 | 12.7 |
| Penfluridol † | 4.5 | 9.0 | 13.5 |
| Pimozide | 3.0 | 6.0 | 9.0 |
| Quetiapine | 274 | 548 | 822 |
| Risperidone | 3.8 | 7.5 | 11.3 |
| Sulpiride | 600 | 1200 | 1800 |
| Trifluoperazine | 10 | 20 | 30 |
| Xanomeline–trospium ‡ | 125 | 250 | 375 |
| Ziprasidone | 56.5 | 113 | 170 |
| Zuclopenthixol | 22.5 | 45 | 67.5 |
| **Clozapine, by sex and smoking (mg/day)** | | | |
| Female, non-smoker | 133 | 265 | 398 |
| Male, non-smoker | 162 | 323 | 485 |
| Female, smoker | 216 | 431 | 647 |
| Male, smoker | 263 | 525 | 788 |
| **Long-acting injectables (mg per stated interval)** | | | |
| Aripiprazole, monthly | 200 | 400 | 600 |
| Aripiprazole, 2-monthly | 480 | 960 | 1440 |
| Paliperidone, monthly | 75 | 150 | 225 |
| Paliperidone, 3-monthly | 263 | 525 | 788 |
| Paliperidone, 6-monthly | 500 | 1000 | 1500 |
| Risperidone, 2-weekly | 25 | 50 | 75 |
| Olanzapine, 2-weekly | 150 | 300 | 450 |
| Olanzapine, 4-weekly | 203 | 405 | 608 |
| Haloperidol decanoate, 4-weekly | 150 | 300 | 450 |
| Flupentixol decanoate, weekly | 200 | 400 | 600 |
| Zuclopenthixol decanoate, weekly | 300 | 600 | 900 |
| Fluphenazine decanoate, 2-weekly | 50 | 100 | 150 |

*The column headed 100 is the AP_Load2026 reference dose of Tables S1 and S2, and, for clozapine, of main-text Table 1, restated as the dose a patient must be prescribed to score 100; the 50 and 150 columns are half and one and a half times that dose, so that the scale can be read in familiar milligrams. Oral doses are mg/day; injectable doses are mg per stated dosing interval, used directly without conversion to a daily equivalent. Clozapine smokers are shown at full induction (≥10 cigarettes/day); intermediate consumption is interpolated, so every dose between the non-smoker and smoker rows is attainable. Because the total is not capped, a score of 150 is meaningful and arises from high-dose treatment, polypharmacy or augmentation; for agents whose reference is the label maximum itself — every injectable and the oral agents with no assigned WHO DDD — it necessarily lies above the licensed maximum, and is shown as an arithmetic property of the scale, not as a recommended dose. † Agents outside the UK formulary (Table S1); penfluridol is dosed weekly and the value shown is its daily equivalent. ‡ Scored on the xanomeline component. Values are rounded for display; scoring uses the unrounded references.*

### Table S9. Consistency of candidate reference-dose systems with external criteria

Consistency of five candidate reference-dose systems with published reference doses derived independently of both the DDD and the licensed maximum. Lower SD of the log ratio indicates a reference that is more consistently proportional to the external criterion across drugs; a uniform offset is unimportant on a ratio scale. The WHO DDD and the flat 1.5 × DDD give identical values by construction, since one is a constant multiple of the other, and both are shown for completeness.

| **Reference system** | **SD log ratio vs Gardner (n=14)** | **SD log ratio vs Leucht ED95 (n=6)** |
| --- | --- | --- |
| Capped geometric mean (adopted) | 0.240 | 0.293 |
| Flat 1.5 × DDD | 0.259 | 0.287 |
| WHO DDD | 0.259 | 0.287 |
| Geometric mean (uncapped) | 0.289 | 0.291 |
| %BNFmax | 0.435 | 0.337 |

### Figure S1. The same prescribed dose scored against three references

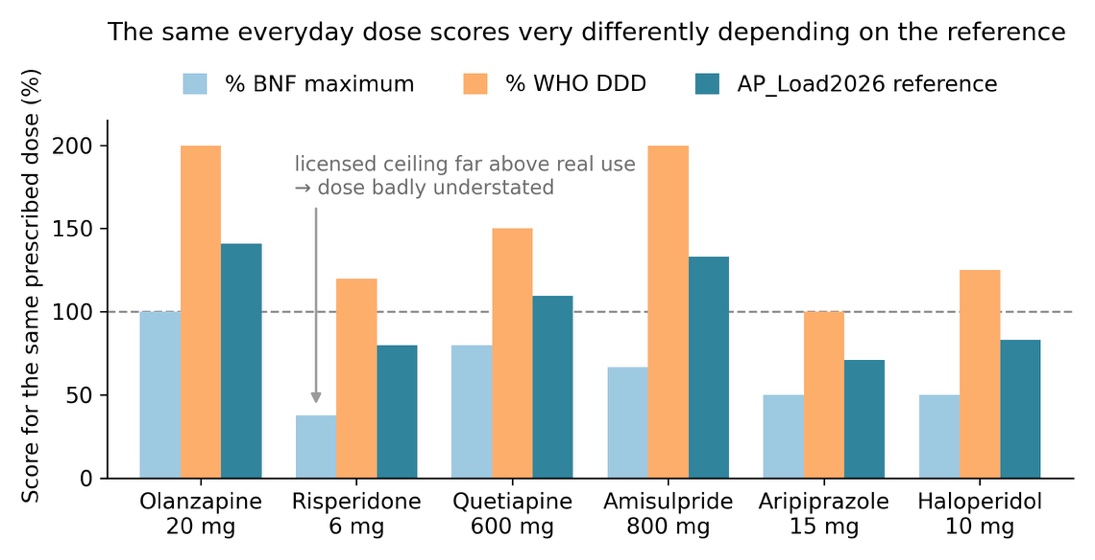

The same prescribed dose scored against three references. Values above 100% indicate a dose exceeding the reference. The licensed maximum understates agents such as risperidone whose ceiling sits far above the doses actually used, whereas the DDD over-weights ordinary doses. Only non-clozapine agents are shown, since the clozapine reference is individualised; clozapine is compared across the four sex–smoking strata in Table 4 of the main text.

### Figure S2. Sensitivity of the non-clozapine reference to the cap value

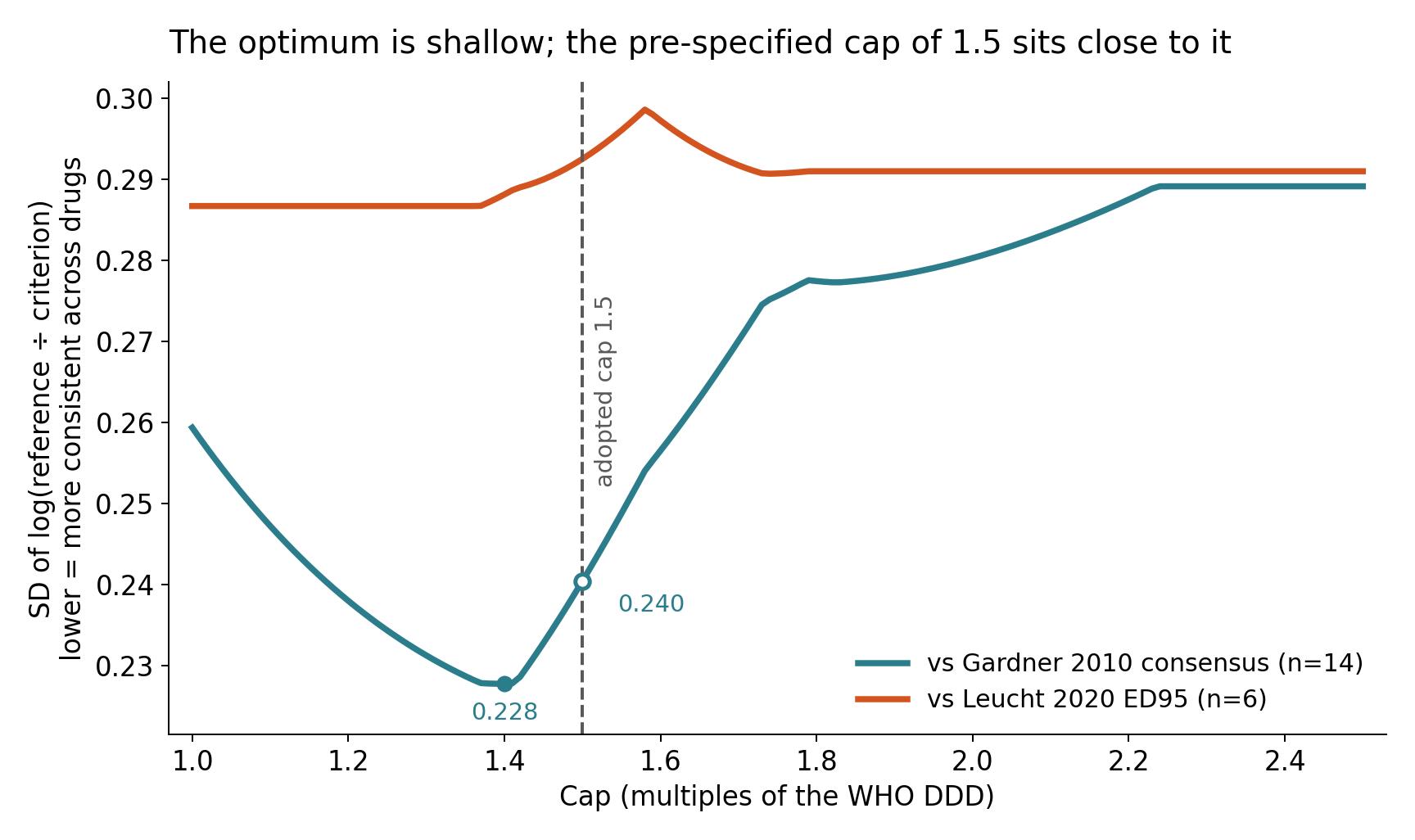

Sensitivity of the non-clozapine reference to the cap value. The optimum against the Gardner criterion is shallow: the minimum of 0.228 near 1.4 differs little from the 0.240 obtained at the adopted value of 1.5, and the difference is small relative to sampling variation across fourteen drugs. The Leucht criterion shows no discernible optimum.
